# Attitude and Behavior of People Towards Visiting the Hospital During the Pandemic

**DOI:** 10.1101/2021.08.21.21261953

**Authors:** Manaswi shamsundar, Shaista Choudhary

## Abstract

**Introduction:** Visiting the hospital is important as a part of check-up in avoiding the major risks of unknown and serious diseases irrespective of any pandemic. Our study aims to understand the attitude and behaviour of common people towards visiting the hospital during COVID-19 pandemic in two major hotspot states of India namely Maharashtra and Karnataka.

**Methodology:** A cross-sectional study was conducted between July-august 2021 among the population of two states. A total of 636 respondents completed the survey and returned electronically. Data were analysed using suitable statistical tools to achieve the objective of the study.

**Results:** 74.8% of the respondents were not ready to visit the hospitals during the COVID-19 pandemic unless the symptoms were serious. On the other hand, 25.2% of the of respondents were willing to go to the hospitals. The top three reasons for the reduction in visits are fear of getting infected in the hospitals by COVID-19 patients (72.6%), fear of stepping out of home (31.1%) and fear of COVID-19 infection by the lab equipment (24.5%).

**Conclusion:** Overall the study revealed that there was a reduction in number of visits to the hospitals for common diseases in people after the pandemic started. But the people were still willing to go to the hospitals if they noticed any major symptoms or symptoms related to COVID-19. Our findings may be useful to develop strategies to address concerns in order to ensure that people don’t get any serious illness because of the fear of COVID-19.

## 1. Introduction

Coronavirus disease 2019 (COVID-19) is caused by severe acute respiratory syndrome coronavirus 2 (SARS-CoV-2). In December 2019, the COVID-19 pandemic outbreak occurred in Wuhan China, and spread out rapidly around the world. As of 1^st^ August 2021, more than 196.9 million cases have been reported across 213 countries and territories with more than 4.2 million deaths and more than 1135.19 million people have been fully vaccinated. In India, as of 1^st^ August, 2021 more than 31.6 million cases have been reported more than 0.42 million people have died and more than 101.6 million have been fully vaccinated. (1)

Fig. 1(a) shows statistics of the COVID 19 across the world and Fig 1(b) shows statistics of COVID-19 in India.

**Fig. 1.**
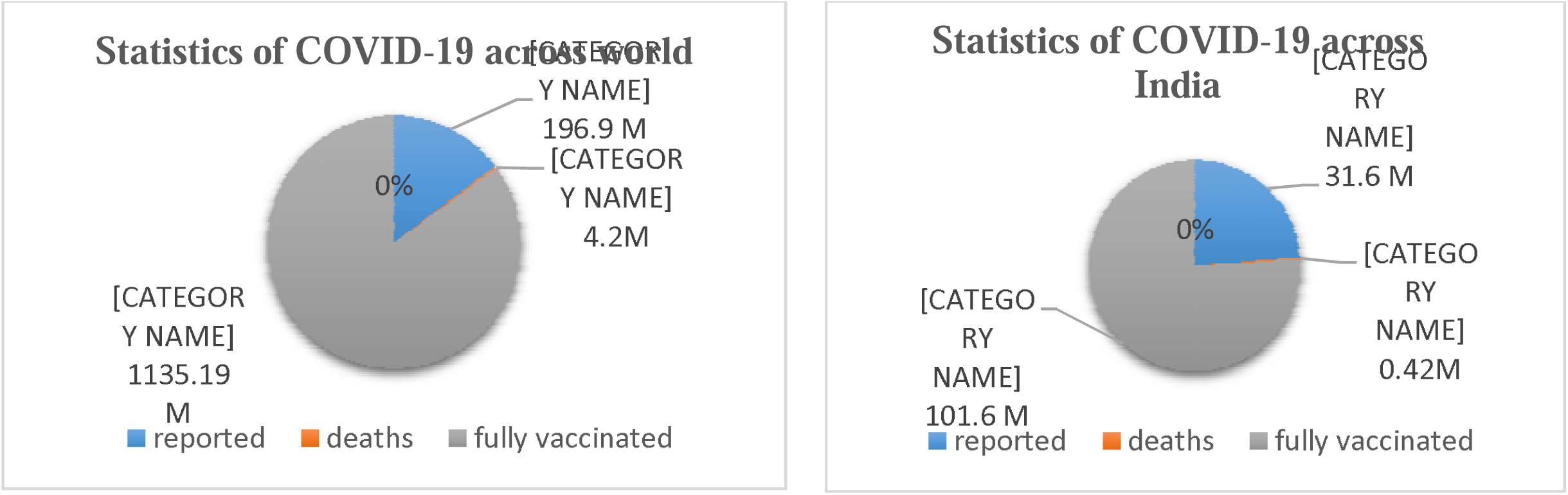
(a) Statistic of the COVID 19 across world Fig. 1(b) Statistic of the COVID 19 across India

Visiting the hospitals has become a routine approach of people; some visit routinely for their long term diseases like diabetes mellitus, allergies, heart disease, thyroid’s disease, blood pressure, etc. and some of the people visit the hospitals even for some illnesses like cold and flu, headache, burns, cuts, etc. One of the major reason of why people visit the hospitals is the fear of possible diagnostic delays in life-threatening or crippling diseases. Visiting the hospital has become a part of life, but after the attack of the COVID-19 pandemic, it has been observed a serious reduction in people visiting the hospitals, or even the health care workers avoiding patients visiting the hospitals. Thus the pandemic has indirectly affected access to health care for patients with other diseases which are not related to COVID-19. This situation has also been observed in some of the countries where the pandemic is not yet overwhelmed by an outbreak. In some countries it is realized that other patients are being affected in an indirect pathway and rebound in visits has occurred across all specialties. (2) Nevertheless, the relative decline in visits remains largest among surgical and procedural specialties and paediatrics. The relative decline is least in other specialties such as behavioural health and adult primary care. The rebound is smaller among school children and larger among older adults. (3) A major reduction in the Emergency room (ER) visits of patients in most of the hospitals during the pandemic period is due to social distancing and country “lock-down”. (2) This reduction of visits to the hospitals may cause the building up of the disease-causing agents in the human body and may lead to major problems with multiple diseases in a long way.

The present study presents insights regarding the attitude and behaviour of common people towards visiting the hospitals in India only for Maharashtra and Karnataka which are the major hotspot states in India.

The study includes to find out how people respond to the ongoing pandemic situation, their willingness to visit the hospitals after the pandemic started, to know the reduction of number of visits and the main reasons to avoid the hospital visits. These major concerns are addressed among the other concerns to reduce the risk of other health-related deaths during the pandemic.

Therefore, our study aims to ask and find out the answers to all these major concerns and to address them properly. A major strength of our study is the large sample size, representation from different genders and age groups. The survey questionnaire was available in English and distributed in an online format, which can further introduce selection bias favouring English-literate people and those with access to the Internet.

## 2. Methodology

### 2.1 Study Design and Sampling Technique

This is a cross-sectional survey among common people in two states of India (Maharashtra and Karnataka). Conventional non-probability sampling technique was used to analyse the questions given to the respondents. (4) The inclusion criteria were common people (undergraduate, postgraduate, and the people from medical field) at the time of data collection and having access to an internet connection to fill out the online questionnaire. Individuals who did not fill the form completely were excluded from the study.

### 2.1 Study Instrument and Administration

A short online questionnaire was developed after a review of a similar study. (2) It comprises demographic characteristics of the respondents such as; age, sex, nationality, profession. (4) Outcome variables include the respondent’s attitude towards visiting the hospitals and the reasons for not visiting the hospitals during the COVID-19 pandemic. The following ten questions were formulated for the online survey through google forms as shown in Table 1. The electronic questionnaire was generated and entered into an online survey system using google forms. The link to respondents was distributed across social media platforms and the data collection took place in June 2021. The typical Google form is shown in Fig 2.

**Table 1.** Electronic questionnaire generated for online survey.

| **Question No.** | **Question** | **Answer options** |
| --- | --- | --- |
| 1. | Were you going to the hospital for common illness before the pandemic started? | yes or no |
| 2. | Will you go to the hospital for illness having covid-19 related symptoms after the pandemic has started? | yes or no |
| 3. | Are you going to the hospital for other common health problems related to eye ear skin or joints after the pandemic has started? | yes or no |
| 4. | Are you under treatment of any long term diseases (like Diabetes or BP)? | Yes or no |
| 5. | Has the frequency of you visiting the hospital reduced after the pandemic started? | yes or no |
| 6. | How often do you go to the hospital after the pandemic started? | with frequency given |
| 7. | What is the reason behind reducing the hospital or lab visits? | With options given and further if any, asked to type the answers |
| 8. | If you are reducing the number of visits how are you managing your health problem? | With options given and further if any, asked to type the answers |
| 9. | Safety measures you take when you visit a the hospital or lab. | With options given and further if any, asked to type the answers |
| 10. | Has your frequency of visits to the hospital changed after taking vaccine? | yes or no |

**Fig 2.**
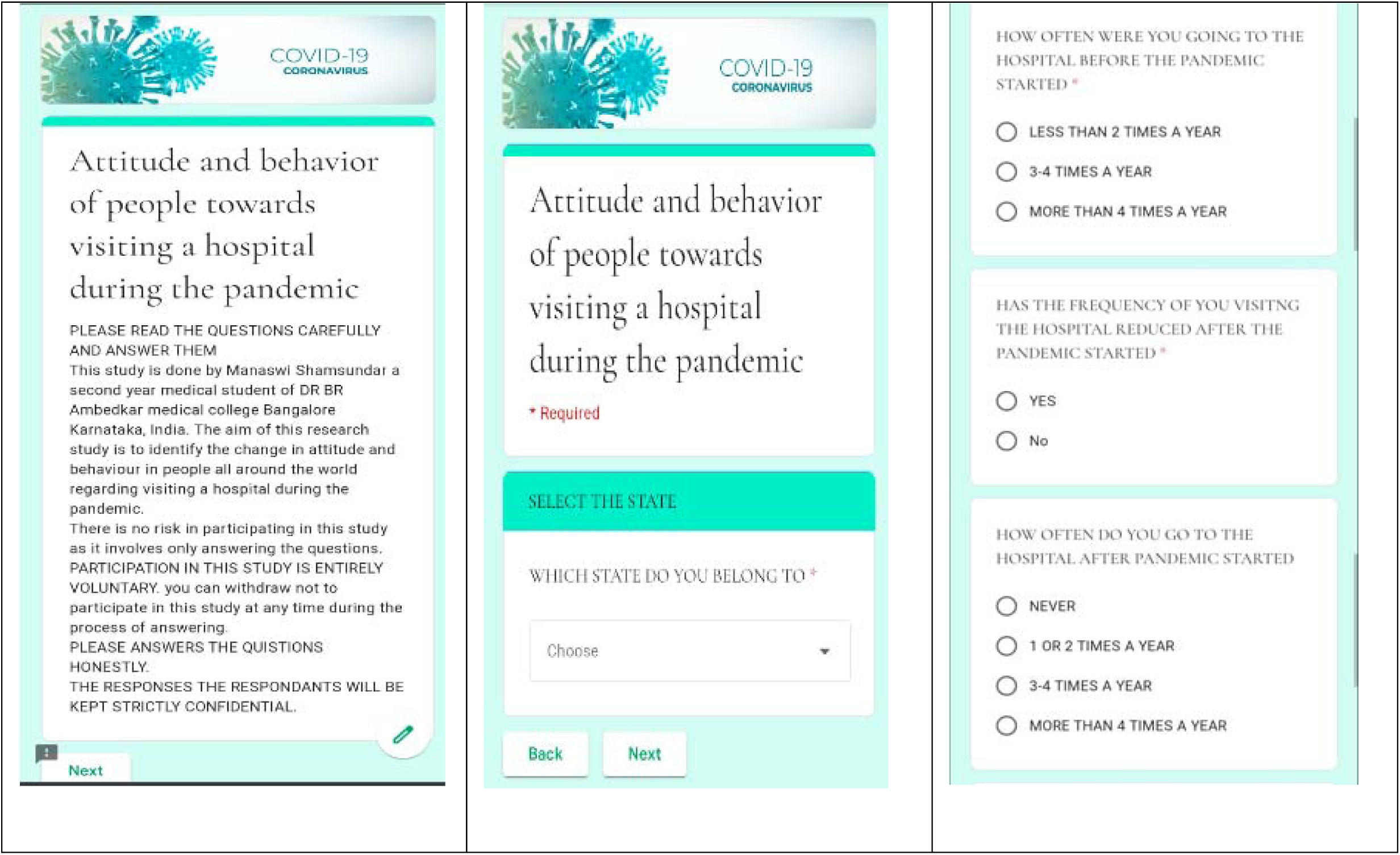
A typical google form.

### 2.2 Data Analysis

There were around 636 respondents representing females and males from two states of India who responded to the survey through google forms. Google forms data were downloaded and data were analysed using different statistical tools. The tools are being used to verify the normal distribution of variables and comparisons between groups for categorical variables. Binary logistic regression analysis was carried out to identify parameters more strongly associated with respondent’s Attitudes and behavior of people towards visiting the hospitals during the pandemic.

### 2.3 Ethical Considerations

Anonymity and confidentiality was ensured by not attaching any names or identifiable codes to the online questionnaires and the rights of the participants to withdraw from the study anytime was also clearly stated in the online survey form.

## 3. Results

### Distribution and sociodemographic characteristics of respondents

A total of 1590 respondents representing females (48.4%) and males (50.3%) from two states of India completed the online survey. The mean age of participants was 39.9 years. There were 37.4% respondents were postgraduate & above and 35.8% respondents were undergraduate and remaining respondents were high school graduates and college graduates. Among 1590 of respondents 30.1 % of respondents are from the medical field. Table 2 shows the distribution of sociodemographic characteristics used in the study.

**Table 2.** Distribution of Sociodemographic Characteristics.

| **Socio demographic data** | **Number** | **Percentage** |
| --- | --- | --- |
| **States** |  |  |
| Maharashtra | 352 | 55.3% |
| Karnataka | 264 | 41.0% |
| others | 20 | 3% |
| **Age** |  |  |
| 18-45 | 419 | 65.8% |
| 46-70 | 199 | 31.2% |
| >70 | 18 | 4.9% |
| **Gender** |  |  |
| Male | 328 | 50.5% |
| female | 308 | 48.4% |
| **Profession** |  |  |
| Postgraduate and above | 238 | 37.4% |
| Undergraduate | 228 | 35.8% |
| College | 154 | 24.2% |
| High school | 16 | 2.5% |
| People from medical field(among total 636 of respondents) | 192 | 30.1% |

The analysis of each questionnaire are as follows:

### 3.1 Were you going to the hospital for common illness before the pandemic started?

More than half of respondents i.e. about 62.6% were visiting the hospital for common illnesses like the common cold, cough headache, body ache, first degree burns, acidity, indigestion, eye, ear, joint, or skin related problems, etc. before the pandemic started. On the other hand, 37.1% of respondents were not visiting the hospitals for such common illnesses even before the pandemic started. Fig. 3 shows the percentage of people visiting to the hospital before the pandemic started.

**Fig. 3.**
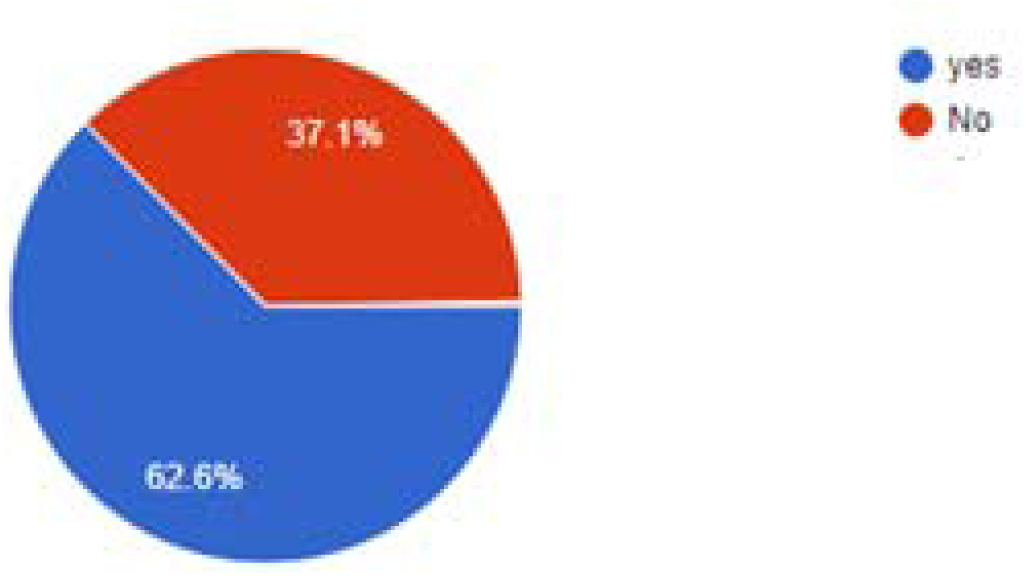
Percentage of people visiting to the hospital before the pandemic started.

### 3.2 Will you go to the hospital for illness having COVID-19 related symptoms after the pandemic has started?

More than half of respondents i.e. about 67% are visiting the hospital for illness having COVID-19 related symptoms like the common cold, cough headache, body ache which lasted for a long time after the pandemic started. On the other hand, 32.7% of respondents were not visiting the hospitals for illness even though they are having covid-19 related symptoms after the pandemic started. Fig. 4 shows the percentage of people visiting the hospital after the pandemic started.

**Fig. 4.**
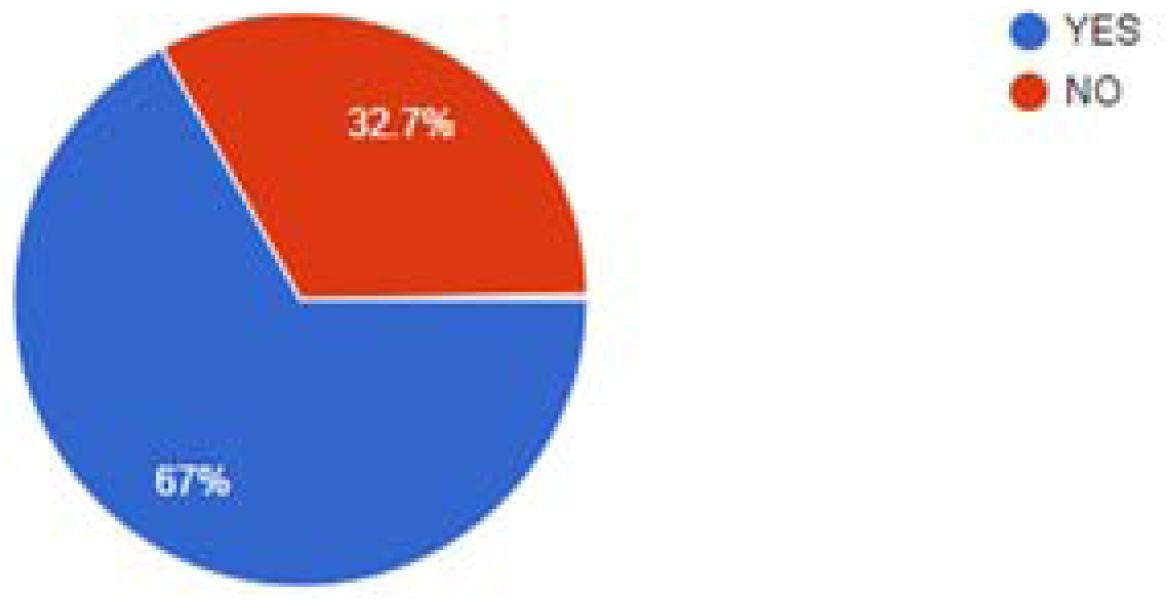
Percentage of people visiting to the hospital after the pandemic started.

### 3.3 Are you going to the hospital for other common health problems related to eye, ear, skin or joints after the pandemic has started?

More than half of respondents i.e. 60.9% are not visiting the hospital for other common health problems like first degree burns, acidity, indigestion, eye, ear, joint, or skin -related problems after the pandemic has started. On the other hand, 39.1% of respondents were visiting the hospitals for other common health problems after the pandemic started. Fig. 5 shows the percentage of people visiting the hospital after the pandemic started.

**Fig. 5.**
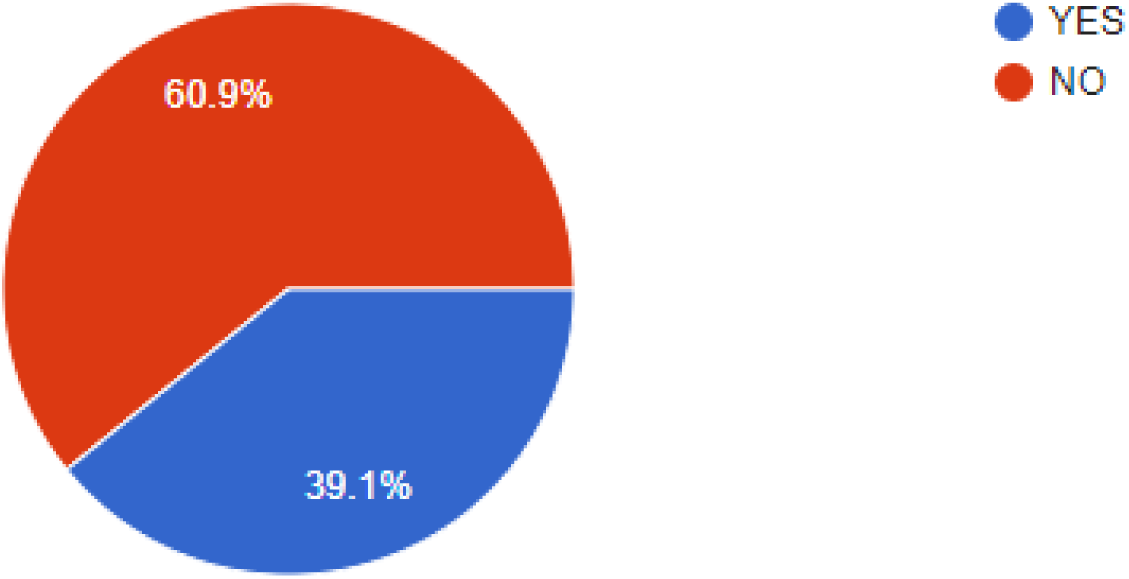
Percentage of people visiting to the hospital after the pandemic started.

### 3.4 Are you under treatment of any long term diseases (like Diabetes or BP)?

About 86.5% of respondents were not under any long-term treatment because of the average age group of respondent being 39.9 years, on the other hand, 13.5% of respondents were under the treatment of long term disease. This question was asked to know how people are maintaining the long term diseases like diabetes or blood pressure (commonly) during the COVID-19 pandemic when they are afraid to go out for regular follow-ups and laboratory tests. Fig. 6 shows the percentage of people having long term diseases.

**Fig. 6.**
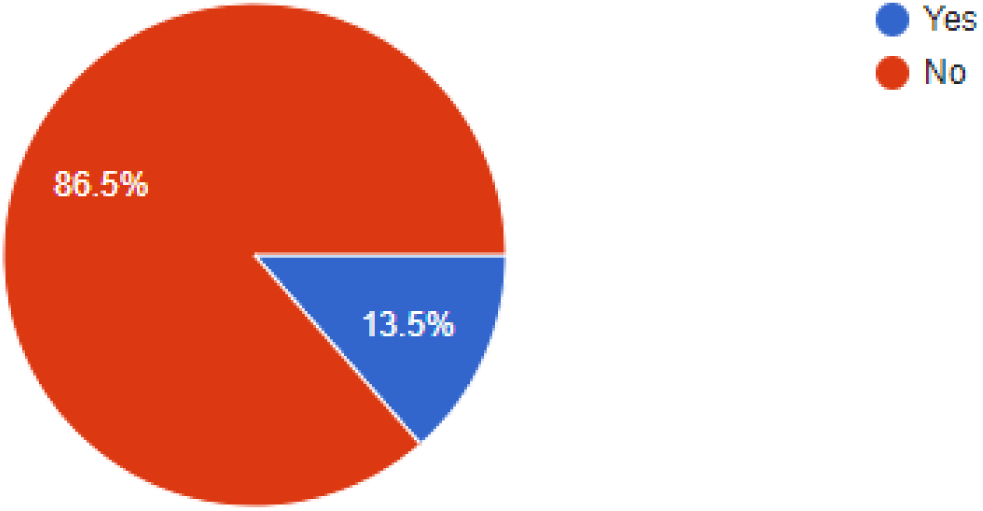
Percentage of people having long term diseases.

### 3.5 How often were you going to the hospital before the pandemic started?

About 73% of respondents are visiting the hospital less than two times a year, 19.8% of respondents are visiting three-four times a year and 7.2% of respondents are visiting more than four times a year for any disease before pandemic started. Fig. 7 shows the percentage of visits to the hospital before the pandemic started.

**Fig. 7.**
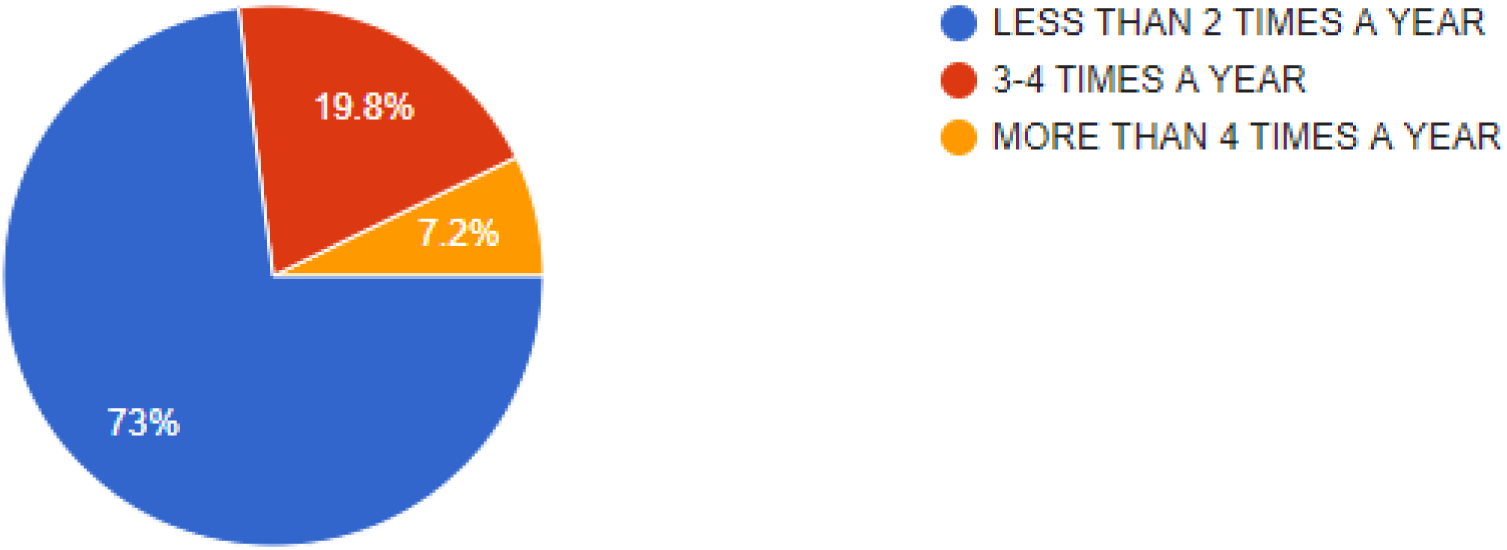
Percentage of visits to the hospital before the pandemic started.

### 3.6 Has the frequency of you visiting the hospital reduced after the pandemic started?

About 74.8% of respondents stated that the frequency of visiting the hospital has reduced after the pandemic started and 25.2% of respondents stated that the frequency of visiting the hospital has not reduced even after the pandemic started. Fig. 8 shows the percentage of people who have reduced the number of visits to the hospital after the pandemic started.

**Fig. 8.**
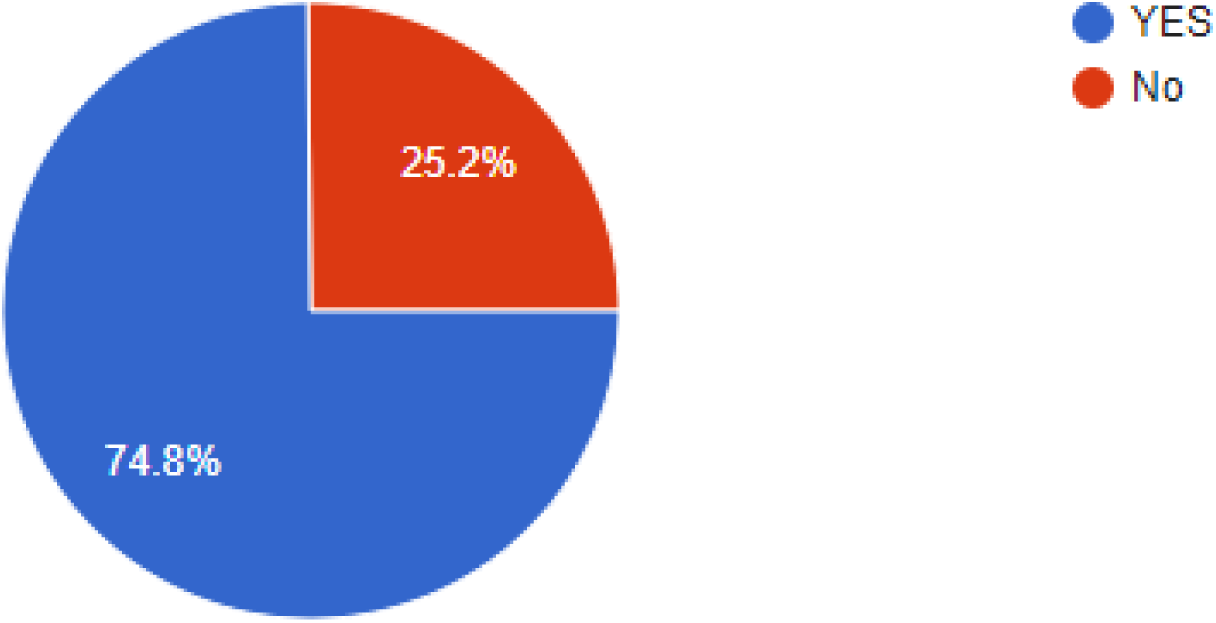
Percentage of people who have reduced the number of visits to the hospital after the pandemic started.

### 3.7 How often do you go to the hospital after the pandemic started?

About 49.7% of respondents are visiting the hospital once or twice a year for any disease, 5.3% of respondents are visiting three-four times a year to the hospitals, 40.6% of the respondents have not visited the hospital since the pandemic has started and other 3.8% of the respondents visit more than four times a year to the hospital. Fig. 9 shows the percentage of visits a person makes to the hospital after the pandemic started.

**Fig. 9.**
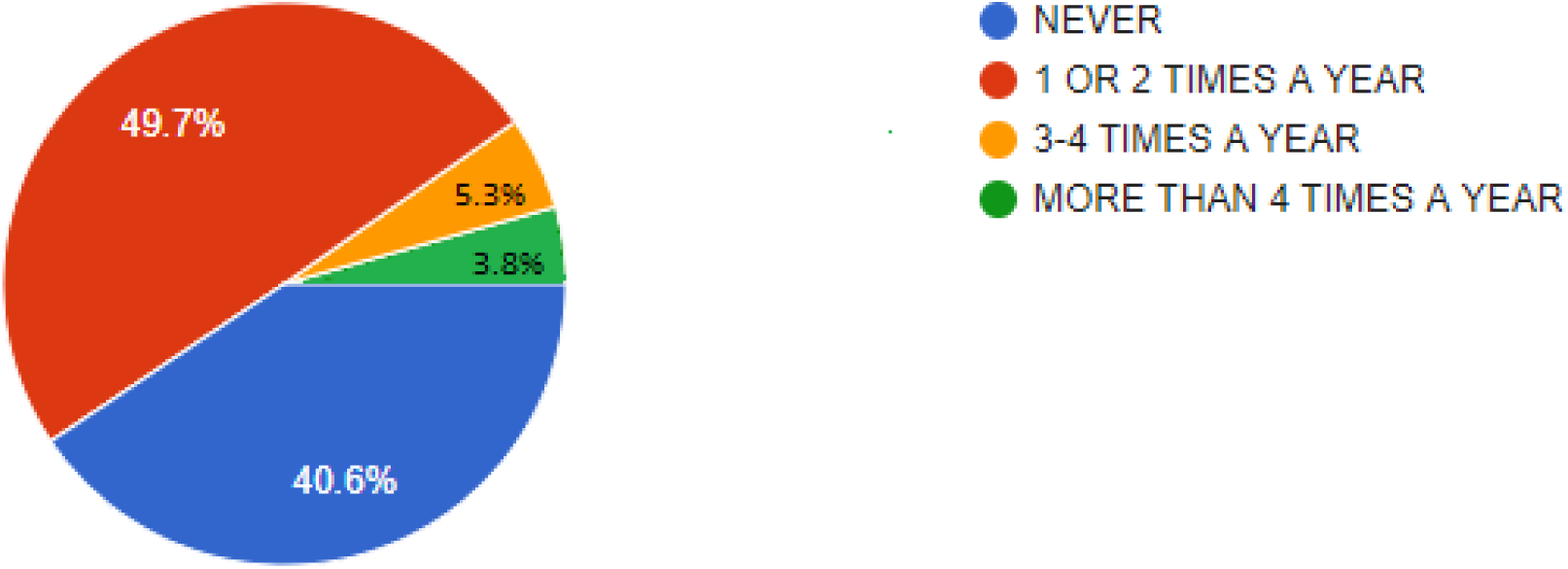
Percentage of visits to the hospital after the pandemic started.

### 3.8 What is the reason behind reducing the hospital or lab visits?

About 59.7% of respondents have reduced the frequency of visiting to the hospital during the pandemic due to the fear of getting infected by coronavirus in the hospital, 22.6% of respondents due to fear of getting infected by coronavirus outside the hospital, 15.7% of respondents due to fear of getting infected by equipment used in the hospitals, other reasons were lockdown in the city, distrust and misconception, no social distancing in crowded places. Fig 10 shows the major reasons for reduction of visits to the hospital.

**Fig 10.**
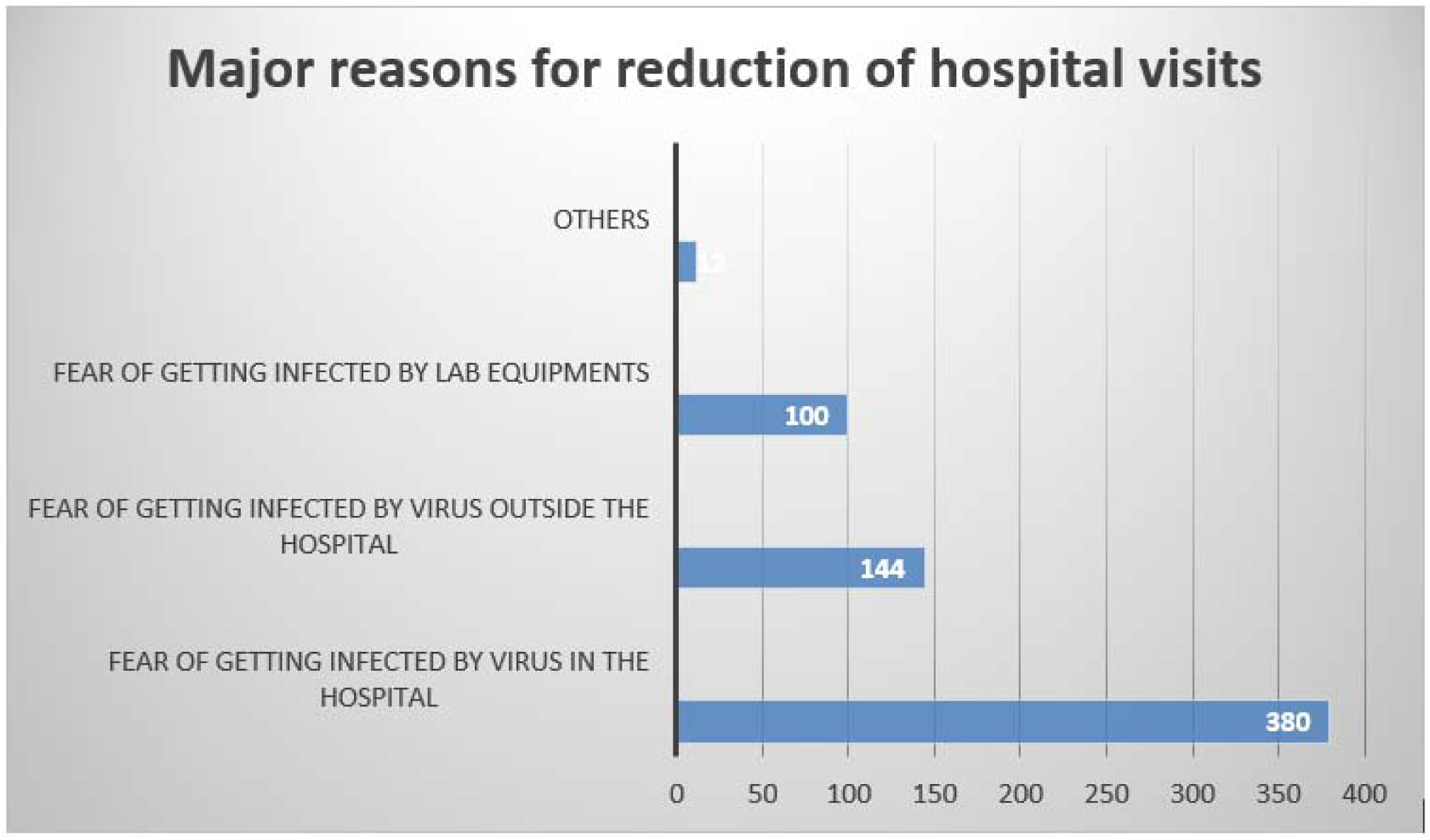
Major reasons for reduction of visits to the hospital.

### 3.9 If you are reducing the number of visits how are you managing your health problem

About 58.4% of respondents are managing their health problems by online consultation of doctors, 20.1% of respondents by taking medications on their own, 19.4% of respondents by calling the doctor home for consultation or lab technician to collect samples if needed. Whereas other respondents are managing by home remedies like homeopathy, yoga, Ayurveda medicines, and immunity boosters. But Some of the patients who have long term diseases go to the hospital for regular follow-ups to avoid unavoidable risks. Fig 11 shows the distribution of management of health when people reduced visiting the hospitals

**Fig 11.**
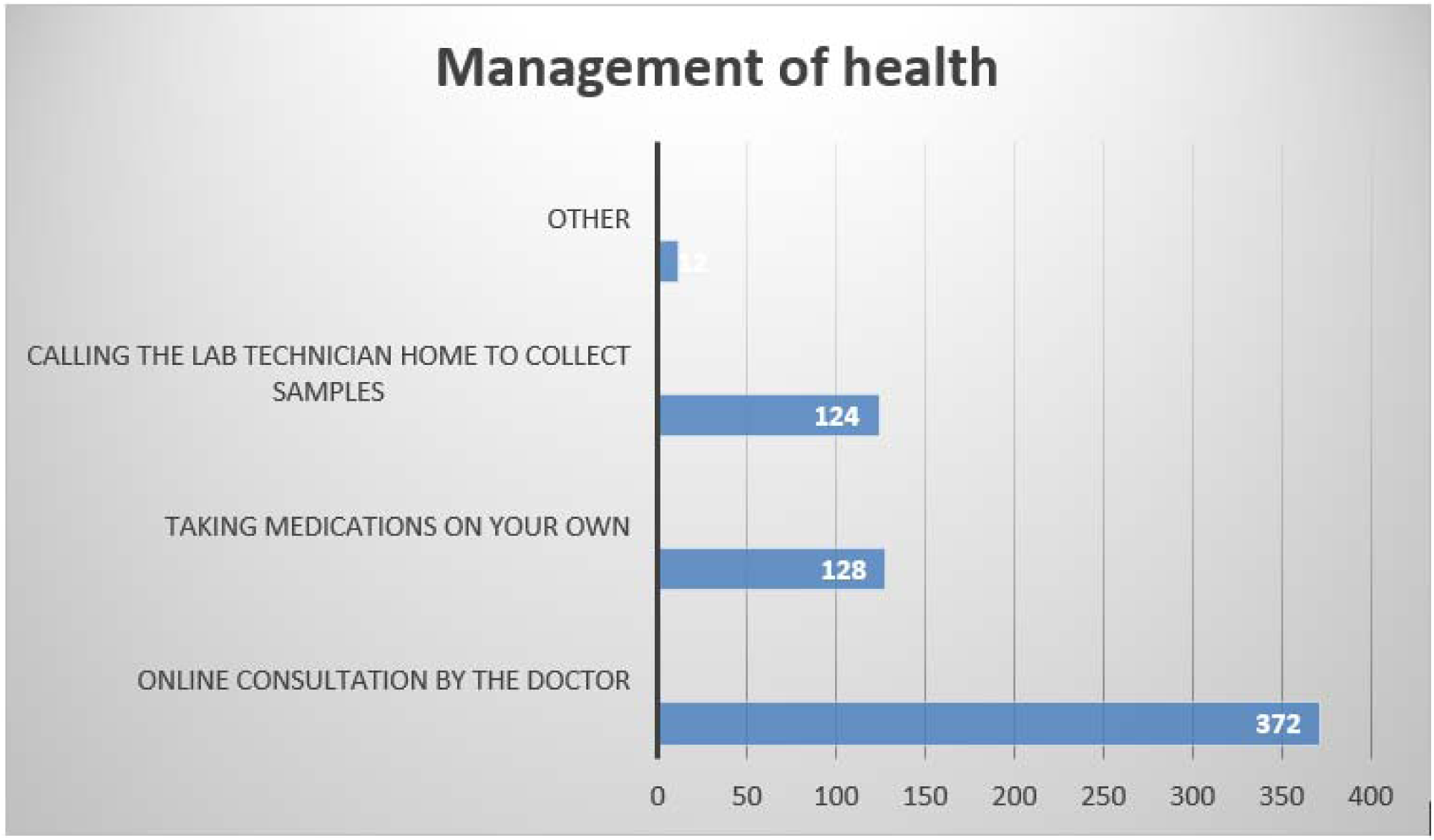
Distribution of management of health when people reduced visiting the hospitals.

### 3.10 Has your frequency of visits to the hospital changed after taking vaccine?

About 53.7% of respondants have not increased the frequency of visiting the hospitals even after taking both the doses of vaccination, on the other hand 27.3% of respondents have gained the confidence to visit the hospital for the treatment after vaccination. Fig. 12 shows the distribution of frequency of visits after taking vaccination.

**Fig. 12.**
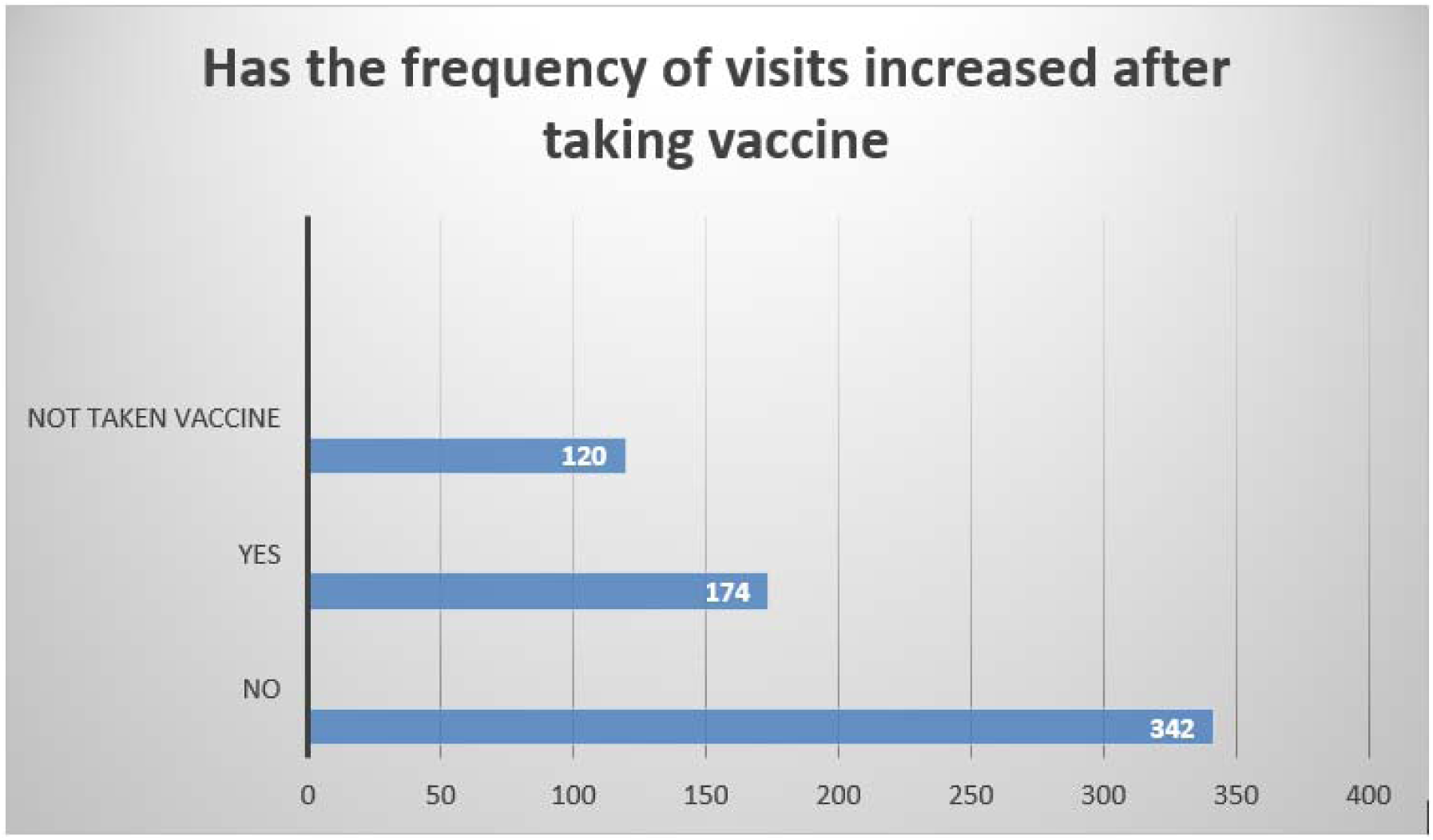
Distribution of frequency of visits after taking vaccination.

## 4. Discussion

Total of 636 respondent’s forms data was analysed. The analysis observes that before the pandemic started around 62.6 % of respondents were visiting the hospitals. The main reason for this is that people are not ready to take any risk for their lives and are afraid that a simple disease may turn into a dangerous disease if they do not visit the hospital.

It is observed that about 67.3 % of people are visiting the hospital for illnesses having COVID-19 related symptoms whereas about 32.7% are not visiting the hospital for any other common health problems after the pandemic started.

The analysis also shows that 73 % of respondents are visiting to the hospital less than two times a year for any disease before the pandemic started but after the pandemic started only 49.7% of respondents are visiting twice a year. Data also shows that about 74.8 % of the respondents reduced visiting the hospital after the pandemic started.

The analysis also shows that 86.5% of respondents are not under any long-term treatment like diabetes or blood pressure this maybe be due to the mean age group of the respondent being 39.9 years. Long-term diseases increase the severity and mortality of COVID-19, thus it is suggested that care for patients with long-term diseases must be increased in order to reduce any further complications and the risk of death. (5,6)

The analysis also shows that the major reasons for the reduction in visits to hospitals is due to getting infected within the hospitals 59.7%, outside the hospitals 22.6%, and due to hospital equipment 15.7%. This analysis reveals that more percentage of respondents have reduced the visit to the hospitals due to getting infected within the hospitals.

The data shows that 58.4% of respondents manage their health themselves by online consultation of doctors, 20.1% by taking medications on their own, 19.4% by calling the doctor or lab technician home for sample collection and others by home remedies. This shows us that people are finding new ways to not to go to the hospital and getting treated at home itself. The majority of the time, the treatment may work but as the doctor cannot see the patient personally, many hidden diagnoses may be left out and the patient may get critically ill because of no early diagnosis.

The analysis reveals that 53.7% of respondents did not increase the frequency of visiting hospitals even after getting fully vaccinated and only 23.3 % of the people gained the confidence to visit the hospitals this reveals that still, the vaccination drive in India is not sufficient to get things back to normal. Patients should be checked about their potential to be expose SARS-CoV-2 in the hospital facility if they are want to visit hospitals even after getting fully vaccinated. (7)

Even hospitals are taking care of the people’s health by reducing the number of patients who visit the hospitals to decrease the risk of transmitting the virus to other patients or health care workers within their practice and to restrict visiting hours to ensure a restful environment for patients and allow clinical staff to work efficiently. (8) Instead of physical visits to hospitals, provision is made to consult through telephone or other virtual platforms. The number of online consultations has increased for upper respiratory tract infection, psychological conditions, COVID-19-related investigations and interventions. The increased online consultations have increased demand for the relevant clinical services and reduced hospital visits, thus decreasing COVID-19 spread. Online consultation is not so useful for people in India as it is moderately limited to only the people who have access to internet or good network for mobile calls, this makes the doctors difficult to diagnose such patients and unfortunately these people have to visit hospitals unwillingly, thus it has also increased the risk of transmitting COVID-19 infection. (9)

The health care centres are therefore increasing the trust of patients towards hospitals by following some safety measures before patients enter the hospitals by using Plexiglas dividers between patients and hospital receptionists, temperature checks of patients, using disinfectant wipes, mandatory wearing of masks, etc. Based on the condition of the patient preliminary health care centres are advising patients not to visit the hospitals if the patients are having any mild symptoms, they are advised to go for telehealth service instead of a physical visit to the hospital and to visit hospitals only in case of emergency situations with all necessary precaution.(10) Further evidence is emerging that people instead of visiting hospitals and get further treatment and speedy recovery they socially isolate themselves from family members, which may indirectly cause psychological stress, delay in recovery, mental health problems.(11) Instead of increasing stress of some patients at home with the fear of isolation and family member’s misconception towards the disease, it’s better to visit the hospital and get treated, this will reduce the serious mental health problems of patients. It is clear from psychiatrists and allied professionals, that the COVID-19 pandemic has led to a response that mental health should clearly be taken into consideration at multiple levels – in the general population, among healthcare workers, and in vulnerable populations.(12)It is seen that with increased public reporting based on patient ‘s satisfaction and efforts to improve patient and family engagement, hospitals may consider decreasing their current restrictions on patient visits as, liberal visitation practices can decrease patient anxiety and benefit patients and families.(8)

Hence the study reports that an authority has to design some steps to manage the pandemic inside the hospitals and labs. This data analysis emphasizes the need for adequate management towards non-COVID-19 diseases and their impact on patient’s health during the pandemic.

## 5. Conclusion

This study reveals that there is drastic reduction in hospital visits due to a reduction in attitude and behaviour of common people towards visiting the hospital during the COVID-19 pandemic due to the fear of getting an infection by the virus. This study suggests hospitals to develop some strategies to improve the management of the health care and thereby increase people’s trust towards hospitals so that they start visiting the hospitals to avoid the late diagnosis of any life-threatening or crippling diseases.

## Data Availability

All the data are available in the manuscripts

https://news.google.com/covid19/map?hl=en-IN&gl=IN&ceid=IN%3Aen&state=3

## Informed Consent Statement

Informed consent was obtained from all subjects involved in the study.

